# Examining COVID-19 Mortality Rates by Race and Ethnicity Among Incarcerated People in U.S. State Prisons

**DOI:** 10.1101/2022.06.24.22276470

**Authors:** Mimi Yen Li, Shelby Grebbin, Ankita Patil, Tori L. Cowger, Dennis Kunichoff, Justin Feldman, Monik C. Jiménez

**Affiliations:** Harvard Medical School, Boston, MA; Brigham and Women’s Hospital, Boston, MA; Harvard T. H. Chan School of Public Health, Boston, MA

**Keywords:** Incarceration, COVID-19, health disparity

## Abstract

**Objective:** To estimate coronavirus disease 2019 (COVID-19) mortality rates among individuals incarcerated in U.S. state prisons by race and ethnicity (RE).

**Design:** Retrospective population-based analysis

**Setting:** Data from state-level Departments of Corrections (DOCs) from March 1 through October 1, 2020.

**Participants:** Publicly available data collected by Freedom of Information Act requests representing adults in the custody of US state DOCs.

**Main Outcomes:** Cumulative COVID-19 death and custody population data. Crude RE-specific cumulative death rates per 1,000 persons, by state and in aggregate, using RE-specific custody population on March 1, 2020, as the denominator. Rate ratios (RR) and 95% confidence intervals (95%CI) compared state-level and aggregate cumulative age-adjusted mortality rates as of 10/01/2020 by RE, with White individuals as reference group.

**Results:** Of all COVID-related deaths in U.S. prisons through October 2020, 23.35% (272 of 1165) were captured in our analyses. The average age at COVID-19 mortality was 63 years (SD=10 years) and was significantly lower among Black (60 years, SD=11 years) compared to White adults (66 years, SD=10 years; p<0.001). In age-standardized analysis, COVID-19 mortality rates were significantly higher among Black (RR=1.93, 95% CI: 1.25-2.99), Hispanic (RR=1.81, 95% CI: 1.10-2.96) and those of Other racial and ethnic groups (RR=2.60, 95% CI: 1.01-6.67) when compared to White individuals.

**Conclusions:** Age-standardized mortality rates were higher among incarcerated Black, Hispanic and those of Other RE groups compared to their White counterparts. Greater data transparency from all carceral systems is needed to better understand populations at disproportionate risk of COVID-19 morbidity and mortality.

## Introduction

In the United States (U.S.), historically marginalized racial and ethnic (RE) groups, such as Black, Indigenous, and Hispanic/Latinx individuals, have a 2-3 times greater hospitalization risk and nearly 2 times greater mortality risk from coronavirus disease 2019 (COVID-19) compared to White individuals.^1^ Moreover, the COVID-19 mortality rate for Black populations is double that of their White counterparts.^1^ The drivers of these inequities are rooted within historical and contemporary mechanisms of structural racism which result in health-harming exposures through environmental, economic, and social pathways.^2,3^

The U.S. carceral system is one key manifestation of structural racism, resulting in the disproportionate incarceration of historically marginalized RE groups.^4^ Unsurprisingly, incarceration has emerged as a key risk factor for COVID-19 morbidity and mortality. When compared to the general population, those exposed to incarceration exhibit a 5.5-fold higher COVID-19 case rate and 3-fold higher mortality rate.^5–7^ The impact of COVID-19 among incarcerated people is likely to have a disproportionate burden on historically marginalized RE groups, due to their disproportionate rates of incarceration and life-long and intergenerational exposure to structural racism.^8^ However, there is limited data examining RE inequities in COVID-19 metrics among incarcerated populations.^9,10^ Since data reporting from carceral systems is unregulated, metrics disaggregated by RE or other demographics are rarely reported.^11^ Therefore, we estimated the rate of COVID-19 cases and deaths among individuals incarcerated in U.S. state prisons by RE through data obtained from filing Freedom of Information Act (FOIA) requests.

## Methods

### Study Design and Data collection

We performed a retrospective population-based analysis of COVID-19 data from state-level Departments of Corrections (DOCs). Cumulative person-level COVID-19 case, death and custody population data by RE as of March 1 and October 1, 2020, were collected via FOIA requests. Our data were supplemented with publicly available data, for example, from DOC COVID-19 dashboards or from third party organizations, such as the Texas Justice Initiative (https://texasjusticeinitiative.org). For states that responded with sufficient data to calculate mortality rates, we subsequently requested the age distribution of the custody population by RE to enable age standardization. Race and ethnicity data were collected per each DOC’s classification and categorized as Hispanic, and the following non-Hispanic groups: Asian, Native Hawaiian, and Pacific Islander (AANHPI), Black, Native American and Alaskan Native (NAAN), White, Other, and Unknown. Requests were submitted October 1, 2020, and collection concluded on June 1, 2021.

### Statistical Analysis

States were excluded if they only reported Hispanic ethnicity without racial classification, requested fees ≥$500, or did not provide sufficient data to calculate age-standardized rates. Crude data from all states, irrespective of whether sufficient data to age-standardize were provided, are presented in supplemental tables. Our analysis of aggregate mortality data collapsed RE categories to Black, Hispanic, White, and Other due to sample size considerations. We calculated crude RE-specific cumulative death rates per 1,000 persons, by state and in aggregate using each race-specific custody population on March 1, 2020, as the denominator. Rate ratios (RR) were calculated with White individuals as the reference. The mean age at death was compared across categories of RE by one-way analysis of variance, with a Bonferroni correction for pairwise comparisons.

Age-standardized mortality rates were calculated by aggregating deaths and prison populations into age-race-state strata using White individuals as the standard. Because age categories varied by state, we assigned the midpoint of the age range to each age category to facilitate comparability for modeling. For categories without an upper bound (e.g. 65+), we used publicly available data from the Texas prison population to determine a plausible midpoint. We then estimated death counts using a multilevel negative binomial model with random intercepts for states, modeled age as a second-order polynomial, added an indicator variable for race (White as the reference group), and included an offset for the natural log of the population. Due to a lack of age-level case data, age-standardized case rates could not be calculated. Therefore, crude analyses of case data are presented in the supplement only. All analyses were conducted in R (version 4.0.5).

This study was approved by the Mass General Brigham Institutional Review Board.

## Results

Public records requests were filed with all 50 state DOCs. Sufficient data to calculate crude and age standardized mortality rates were obtained from nineteen states and eleven states, respectively **(Figure 1)**. Data accessibility was limited by nuanced public record policies, i.e. handwritten and mailed requests (WV), state residency requirements (TN, AL), Health Insurance Portability and Accountability Act protections (CA), or reviewers denying data availability (**Supplemental Table A**). Six (23%) of responding states required payment ($25.00-$1369.00). State-mandated request fulfillment deadlines (5 days to indefinitely) were sometimes met with an automatic email confirming FOIA receipt. Racial and ethnic classifications were heterogeneous across states (**Supplemental Table A, Supplemental Table B**).

**Figure 1.**
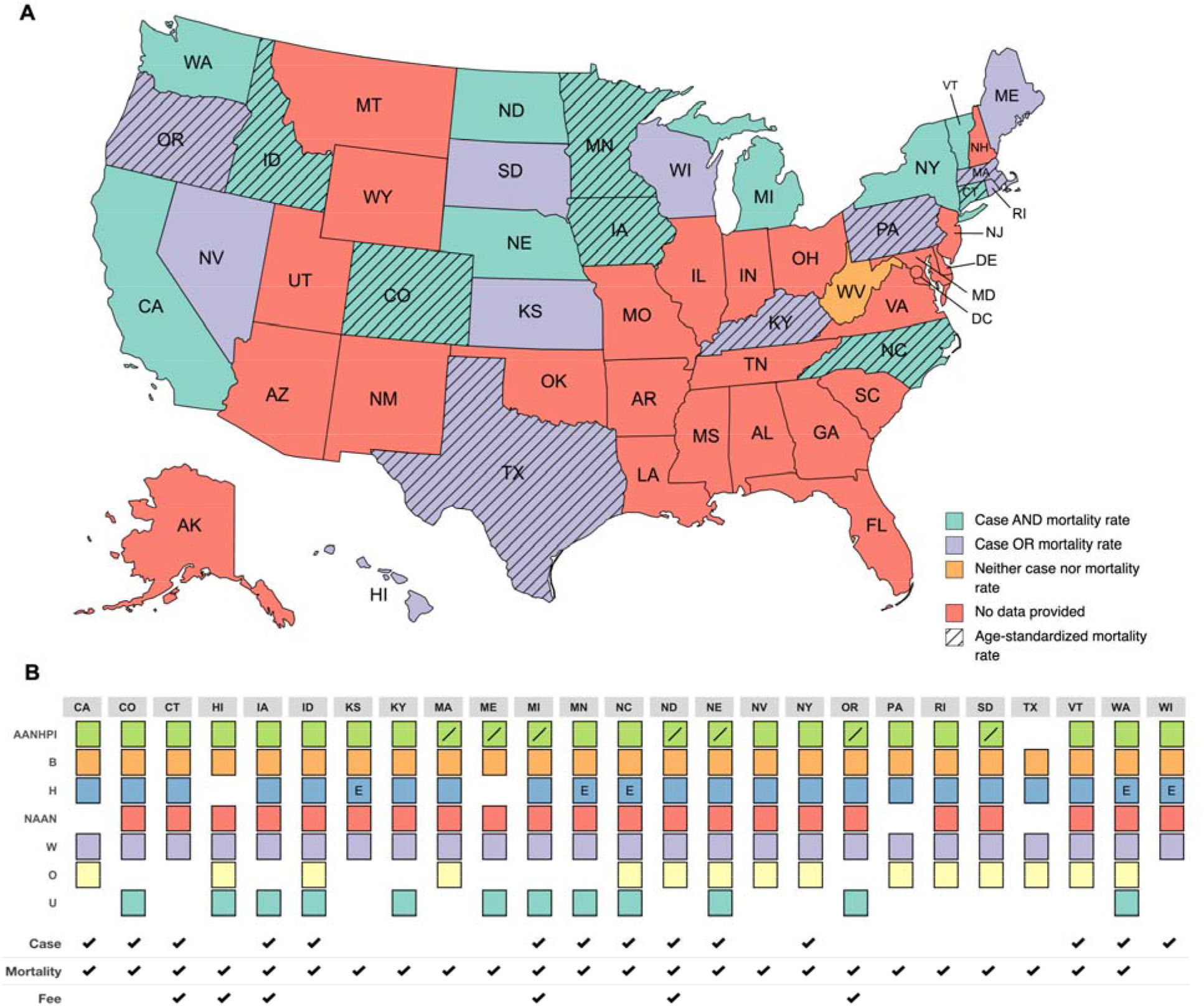
Data, Race, and Ethnicity Reporting from State DOCs. Abbreviations: Asian American, Native Hawaiian, and Pacific Islander (AANHPI), Black (B), Hispanic (H), Native American and Alaskan Native (NAAN), White (W), Other (O), and Unknown (U) (Top) States sending at least partial data are displayed. Figure displays whether each state provided data sufficient to calculate case and mortality rates, case or mortality rates, neither case nor mortality rates, age-standardized mortality rates, or no data at all. States that sent 3/01/2021 custody population, 10/01/2021 case and mortality data by race/ethnicity (all RE groups for case data; White, Black, Hispanic, at minimum for mortality data) were considered having sent all data for case and mortality rate calculations. States that additionally sent mortality data by age are indicated as having sent all data necessary for calculating age-standardized mortality rates. (Bottom) Displays the data sets and racial/ethnic categories reported by each state. States that requested fees are also indicated. States that sent SARS-CoV-2 case (“Case”) and/or mortality data (“Mortality”) by race/ethnicity are marked. Forward slash denotes states that reported Asian American, Native Hawaiian, and Pacific Islander in disaggregated racial categories. “E” denotes states reporting “Hispanic” as an ethnicity rather than a race category.

Mortality data were received from 19 states (CO, CT, IA, ID, KS, KY, MA, MI, MN, NC, ND, NE, NV, NY, OR, PA, RI, SD, VT), however, only 11 states provided sufficient information to calculate age-standardized mortality rates (CO, CT, IA, ID, KY, MA, MN, NC, OR, PA, TX) (**Table 2**).

Our data from 11 states represented 309,273 incarcerated individuals, 58% of whom were classified as Black, Hispanic, or Other RE group. Over the study period, 272 (0.09%) individuals died from COVID-19, of which 94 deaths were among Black adults, 79 among Hispanic, 5 among those of Other RE, and 94 among White adults. Deaths captured in our analyses represent 23.35% (272 of 1165) of COVID-19 deaths in U.S. prisons as of October 1, 2020, based on publicly available datasets.^12^ The average age at COVID-19 mortality was 63 years (SD=10 years) and was significantly lower among Black (60 years, SD=11 years) but not Hispanic (65 years, SD=9) or those of Other RE groups (66 years, SD=8), compared to White adults (66 years, SD=10 years; p<0.001).

Crude mortality rates varied across all states, ranging from 17 per 100,000 people in Colorado to 142 per 100,000 people in Texas **(Table 1)**. In aggregate crude analysis, a significantly greater COVID-19 mortality rate was observed for Hispanic compared to White adults (RR=1.61, 95% CI: 1.18-2.19). However, no statistically significant association was observed among Black (RR=1.24, 95% 0.92-1.66) or those of Other RE (RR=1.12, 95% 0.36-2.71) when compared to White adults.

**Table 1.** Racial and Ethnic COVID-19 Mortality among States with Deaths occurring by October 1, 2020

| **State** | **Black** | **Hispanic\*** | **Other†** | **White** | **n(Deaths) /**<br>**n(Pop)** | **Deaths/**<br>**100,000** |
| --- | --- | --- | --- | --- | --- | --- |
| **CO** | 0/3128 (0.00%) | 1/5588 (0.02%) | 0/908 (0.00%) | 2/7976 (0.03%) | 3/17600 | 17.05 |
| **CT** | 4/5349 (0.07%) | 2/3307 (0.06%) | 0/102 (0.00%) | 1/3653 (0.03%) | 7/12411 | 56.40 |
| **IA** | 1/2128 (0.05%) | 0/580 (0.00%) | 0/235 (0.00%) | 3/5561 (0.05%) | 4/8504 | 47.04 |
| **ID** | 0/260 (0.00%) | 0/1154 (0.00%) | 0/551 (0.00%) | 2/5854 (0.03%) | 2/7819 | 25.58 |
| **KY** | 5/3125 (0.16%) | 0/232 (0.00%) | 0/206 (0.00%) | 6/8639 (0.07%) | 11/12202 | 90.15 |
| **MA‡** | 2/2259 (0.09%) | 0/2064 (0.00%) | 2/266 (0.75%) | 4/3334 (0.12%) | 8/7923 | 100.97 |
| **MN** | 1/3317 (0.03%) | 0/489 (0.00%) | 0/1022 (0.00%) | 1/4101 (0.02%) | 2/8929 | 22.40 |
| **NC** | 9/17805 (0.05%) | 0/1884 (0.00%) | 1/990 (0.1%) | 6/13904 (0.04%) | 16/34583 | 46.27 |
| **OR** | 1/1382 (0.07%) | 2/1948 (0.1%) | 0/735 (0%) | 6/10370 (0.06%) | 9/14435 | 62.35 |
| **PA** | 7/20631 (0.03%) | 1/4313 (0.02%) | 0/368 (0%) | 3/19431 (0.02%) | 11/44743 | 24.58 |
| **TX£** | 64/45761 (0.14%) | 73/46375 (0.16%) | 2/797 (0.25%) | 60/47191 (0.13%) | 199/140124 | 142.02 |
| **Total** | 94/105145 (0.09%) | 79/67934 (0.12%) | 5/6180 (0.08%) | 94/130014 (0.07%) | 272/309273 | 87.95 |

\*Hispanic adults are those of any race, all other groups are non-Hispanic based on data as reported by each state Department of Corrections.
†Includes American Indian, Alaskan Native, Asian American, Native Hawaiian, Pacific Islander, and other racial or ethnic group
‡One individual was reported as American Indian; however, the denominator for American Indian adults in custody was not provided
£Racial and ethnic data from TX are only reported as Black, Hispanic, Other, and White. Therefore, it is assumed that American Indian, Alaskan Native, Asian American, Native Hawaiian, Pacific Islander, are reported in the "Other racial and ethnic" category, but uncertain.

In contrast, in age-standardized analysis, mortality rates were higher among incarcerated Black, Hispanic and those of Other RE groups compared to their White counterparts (**Table 2**). Specifically, the rate of COVID-19 mortality was 93% higher among Black compared to White incarcerated individuals (RR=1.93, 95% CI: 1.25-2.99). Mortality rates were similarly higher among Hispanic compared to White incarcerated individuals (RR=1.81, 95% CI: 1.10-2.96). Mortality rates were also higher among those from Other RE groups, with a 2.60 times higher rate compared to White individuals; however, confidence intervals were wide due to limited sample size (RR=2.60, 95% CI: 1.01-6.67).

**Table 2.** Age-Adjusted Association Between Race and Ethnicity and Coronavirus Disease 2019 Mortality

| Racial/Ethnic Categories | Deaths | Population | Unadjusted |  | Age-Adjusted |  | p-value |
| --- | --- | --- | --- | --- | --- | --- | --- |
|  |  |  | RR | 95% CI | RR | 95% CI |  |
| **Black** | 94 | 105,145 | 1.24 | (0.92-1.66) | 1.93 | (1.25-2.99) | <0.01 |
| **Hispanic\*** | 79 | 67,934 | 1.61 | (1.18-2.19) | 1.81 | (1.10-2.96) | 0.02 |
| **Other<sup>†</sup>** | 5 | 6,180 | 1.11 | (0.36-2.71) | 2.60 | (1.01-6.67) | 0.05 |
| **White** | 94 | 130,014 |  | Ref |  | Ref |  |

RR: rate ratio; 95% CI: 95% confidence interval
\*Hispanic adults are those of any race, all other groups are non-Hispanic based on data as reported by each state Department of Corrections.
<sup>†</sup>Includes American Indian, Alaskan Native, Asian American, Native Hawaiian, Pacific Islander, and other racial or ethnic group

## Discussion

This study is the first systematic analysis of COVID-19 mortality rates by RE across multiple state-level U.S. prisons. Significant RE inequities observed in age-standardized COVID-19 mortality were obscured in crude mortality rate analyses. In our analyses, the rate of mortality in Black and Hispanic incarcerated individuals were found to be 93% and 81% higher, respectively, compared to White incarcerated individuals. Our data demonstrate striking RE inequities in COVID-19 mortality overall and a younger age at death for Black compared to White incarcerated people. These inequities are particularly notable because they persist despite the high case rate among incarcerated individuals overall.^13^

The higher mortality rates of COVID-19 we observed among Black and Hispanic compared to White incarcerated individuals are consistent with and expand upon previous research from Texas prisons.^9^ Between April 1, 2019 and March 31, 2021, the rate of COVID-19 mortality among individuals incarcerated in Texas prisons was significantly higher among Black (RR=1.66, 95% CI: 1.10–2.52) and Hispanic (RR=1.96, 95% CI: 1.32–2.93) compared to White adults in age and sex adjusted analysis.^9^ Our analyses, which examined inequities across 11 state, demonstrate the robustness of such RE inequities and also provide the first estimates for those of Other RE groups. Our analyses therefore provide a broader and more granular understanding of RE inequities across states.

Furthermore, our documentation of high levels of variability and gaps in COVID-19 metric reporting from state carceral systems highlight the significant challenges to characterizing the impact of the pandemic among individuals at the highest risk within our society. The need for intersectional demographic data on COVID-19 is critical and apparent, given our ability to detect significant race and ethnic mortality inequities in age-standardized, but not crude analyses. Lack of adequate data from all state carceral systems masks existing or evolving inequities that may otherwise inform public health measures to prevent the spread of COVID-19, particularly in historically marginalized groups.

Several study limitations should be considered. Our findings of RE inequities are likely underpowered, given our data collection concluded prior to the second surge of COVID-19 in the Fall/Winter 2020.^14^ Yet, our data demonstrate striking RE inequities and a younger age at death for Black incarcerated people compared to their White counterparts, despite the overall young age of incarcerated populations. A lack of complete data from jails, the largest state prison systems (e.g. California and Louisiana), or those with large outbreaks may limit generalizability. Furthermore, standardized joint age and RE distributions were not obtained for all state DOCs, although age-standardization was conducted for 11 of the 19 states with observed deaths and complete RE data. Moreover, the mortality data reported by carceral systems are likely underestimations of deaths directly related to COVID-19 due to limited testing and complications of the disease that were not initially well understood early on in the pandemic.^15^ Further, RE categories were based on state-specific reporting and each DOC’s ascertainment methods were not reported which may lead to misclassification of RE or may mask RE differences. For example, the classification of multiple RE groups under “Unknown,” the aggregation of Asian individuals with Native Hawaiian and Pacific Islander individuals, or Hispanic/Latinos with other RE groups.^16^

### Public Health Implications

Racial and ethnic inequities in COVID-19 mortality among incarcerated populations highlight the need for a more granular understanding of COVID-19’s impact within prison systems. Data transparency and mandated standardized reporting of COVID-19 data by intersectional demographic factors is critical to understanding the full scope of RE inequities in carceral facilities and their impact on the larger community.

Understanding racial and ethnic inequities in carceral systems, one of the key manifestations of structural racism and an extension of our society at large, should be considered in any discussion of the racialized impact of the COVID-19 pandemic.^17,18^ RE inequities among incarcerated individuals are a reflection of the same mechanisms of structural racism that impede optimal health among historically marginalized RE groups in the general population. Disproportionate representation of marginalized RE groups in prisons and high COVID-19 transmissions and deaths, fueled by inhumane confinement conditions, has resulted in significant harm and will continue to perpetuate health injustices over time.

## Data Availability

All data produced in the present study are available upon reasonable request to the authors.

## Ethics approval and consent to participate

Ethical approval was provided by the Partners Massachusetts General Brigham IRB Committee.

## Availability of data and materials

All data generated and datasets used during the current study will be available from the corresponding author on reasonable request.

## Acknowledgements

MCJ is supported by the Brigham and Women’s Hospital Richard Nesson Fellowship. MCJ, JF, DK were supported by The JPB Foundation. Thank you to Karen Jiang, MPH, for her consultation on data analysis and manuscript figure preparation.

## Author contributions

MYL, SG, and MCJ initiated the study, conceptualized the study design, and developed data collection strategy. MYL, SG, AP, MCJ conducted data collection. TC, MCJ, JF, and DK led and conducted the data analysis. MYL, SG, MCJ led and AP contributed to manuscript preparation. All approved the final version of the submitted manuscript.

