## Supplemental Files for "Examining COVID-19 Mortality Rates by Race and Ethnicity Among Incarcerated People in U.S. State Prisons"

### SUPPLEMENTAL MATERIALS

#### Table A. Race and Ethnicity Reporting by State for Reported Data

| State | Date | Metric | Reported Race and Ethnic Categories | Methods Notes |
|---|---|---|---|---|
| AL | AL requires a $25 fee to file a FOIA & informed us they do not collect our requested data | | | |
| AK | No reply after 3 outreach attempts | | | |
| AZ | Requested $1369 for data fulfillment. | | | |
| AR | No reply after 3 outreach attempts. | | | |
| CA | 09/2020 | Population | • Asian, Black, Latino, White, Other<br>• Latino individuals were categorized as Hispanic | CA DOC refused case and death data by RE; Native American or Pacific Islanders reported as "Other." |
| | 12/2019 | Population | • Asian, Black, Hispanic, Native American, Pacific Islander, Other | STATNews |
| | 08/15/2020 | Death | • Asian, Black, Hispanic, Native American, Pacific Islander, Other | STATNews |
| CO | 12/31/2019 | Population | • African American, Asian, Caucasian, Hispanic, Native American | Assumed 0 individuals had Unknown race since not reported as a racial category |
| | 10/01/2020 | Population | • Asian, Black or African American, Caucasian, Hawaiian/Pacific Islander, Hispanic/Latino, Native American/American Indian, Unknown | |
| | 10/01/2020 | Death | • Hispanic/Latino, Caucasian | 0 deaths assumed for all other RE groups |
| CT | 03/01/2020<br>10/14/2020 | Population | • Asian, Black or African American, Hispanic, Native American/ American Indian/Alaska Native, White, None Specified, Other, Unknown | $100.00 fee requested to fulfill request. |
| | 10/14/2020 | Death | • Black, Hispanic, White | 0 deaths assumed for all other RE groups |
| DE | Request denied and we were informed only Delaware residents may file FOIAs in Delaware | | | |
| FL | Request unanswered; redirected to check the Florida DOC website, though this does not provide the data we requested | | | |
| GA | Requested data not available from GA DOC | | | |

| | | | | |
|---|---|---|---|---|
| HI | 03/01/2020<br>10/01/2020 | Population | • Asian or Pacific Islander, Black or African American, Hispanic, Native American/American Indian/Alaska Native, White, None Specified, Other, and Unknown | • $30 paid to file; DOC requested $1,000<br>• DOC indicated that they could not provide case data by race/ethnicity.<br>• 0 deaths were reported for all RE groups |
| IA | 03/01/2020<br>10/01/2020 | Population | • American Indian/Alaska Native, Asian or Pacific Islander, Black, Hispanic, White, and Unknown | |
| | 10/01/2020 | Case | • American Indian or Alaska Native, Asian or Pacific Islander, Black, Hispanic, White | |
| | 10/01/2020 | Death | • Black, White | 0 deaths assumed for all other RE groups |
| ID | 03/01/2020<br>10/01/2020 | Population, case | • Asian, Black, Native American, White, Other, Two or more races, Unknown and Unspecified<br>• *Ethnic Categories:* Hispanic, Not Hispanic, Unknown | |
| | 10/01/2020 | Death | • White | 0 deaths assumed for all other RE groups |
| IL | IL DOC reports they do not maintain records in the manner we requested | | | |
| IN | No response after 3 outreach attempts. | | | |
| KS | FY 2019 | Population | • American Indian, Asian, Black, White<br>• *Ethnic Categories:* Hispanic, Not Hispanic. | KS Fiscal Year 2019 Report |
| | 10/28/2020 | Death | • Black, White | 0 deaths assumed for all other RE groups |
| KY | 03/03/2020<br>10/01/2020 | Population | • American Indian/Alaskan Native, Asian or Pacific Islander, Black, Hispanic/Latino, White, Bi-Racial, Unknown | |
| | 10/01/2020 | Death | • Black, White | 0 deaths assumed for all other RE groups |
| LA | DOC indicated response would be provided but no data received despite follow-up. | | | |
| MA | 03/01/2020<br>10/01/2020 | Population | • American Indian/Alaska Native, Asian, Black, Hispanic, Native Hawaiian or Pacific Islander, White, Other. | |
| | 10/01/2020 | Death | • American Indian, Black, White, Other | 0 deaths assumed for all other RE groups |
| MD | In October 2020, MD DOC requested a 30-day time extension to fulfill request. No data was received from MD DOC. | | | |
| ME | 06/2020 | Population | • Asian, Black or African American, Native American, Native Hawaiian, White, Two or More Races, Unknown | Maine DOC Adult Data Report<br>Reported 0 deaths related to COVID-19. |

| | | | | |
|---|---|---|---|---|
| MI | 03/01/2020<br>09/01/2020 | Population, case data | • American Indian or Alaskan Native, Asian, Black or African American, Hispanic or Latino, Pacific Islander or Native Hawaiian, Unknown, White | |
| | 09/01/2020 | Death | • Black, Hispanic White | 0 deaths assumed for all other RE groups |
| MN | 03/01/2020<br>10/01/2020 | Population, case, death data | • American Indian, Asian, Black, White, Unknown<br>• *Ethnic categories*: Hispanic | |
| MO | Did not fulfill FOIA and redirected us to the MO DOC webpage which does not provide the data we requested. | | | |
| MS | No response after 3 outreach attempts. | | | |
| MT | No response after 3 outreach attempts. | | | |
| NC | 03/01/2020<br>10/01/2020 | Population | • Asian/ORTL, Black, Indian, White, Other, Unknown<br>• *Ethnic categories*: African, American Indian, Asian, European/ N.Am./ Austr, Hispanic/Latino, Oriental, Pacific Islander, Other, Unknown | Race categories were used as race categories and only "Hispanic" was considered ethnicity from the ethnic categories. |
| | 10/01/2020 | Death | • Black, Hispanic/Latino, White, Other | "Hispanic" was reported as race in death data, but 0 COVID-19 deaths in Hispanic individuals. |
| ND | 03/01/2020<br>10/01/2020 | Population | • Asian, Black, Caucasian, Hawaiian, Hispanic, Native American, Other/Mixed | Caucasian and White were aggregated; "Nat," "Native," and "Native American" were aggregated; Hispanic was reported as race in all data, and as ethnicity in case data; counts for Hispanic race and Hispanic ethnicity were the same; 0 deaths from COVID-19 reported/ |
| | 10/01/2020 | Case data | • Asian, Black, Caucasian, His, Nat, Native, White, Other/Multiple<br>• *Ethnic categories*: Hispanic, Not Hispanic | Case data reported at the level of each test, not individual. Records with the same date of birth, sex, race, ethnicity, and facility corresponded to the same individual. |
| NE | 03/01/2020<br>10/08/2020 | Population | • Asian, Black, Hispanic, Native American, Pacific Islander, White, Other, Unknown | 03/01/2020: average daily population 1/1–3/30, 2020; 10/08/2020: average daily population 5/5– 10/8, 2020; Based on reported proportions, we assumed 5 were "Unknown" race and 4 were "Hawaiian and Pacific Islander." |
| | 10/08/2020 | Case data | • Asian, Black, Hispanic, Native American, White, None Specified, Other | |
| | 10/01/2020 | Death data | • Death analyzed as "Unknown" | Death identified by press release |
| NH | NH DOC stated there have been 0 deaths in their system and that they do not track any of the metrics we requested by demographics. | | | |
| NJ | No data disaggregated by race/ethnicity was provided. | | | |

| | | | | |
|---|---|---|---|---|
| NM | No data disaggregated by race/ethnicity was provided. | | | |
| NV | 03/08/20200 9/27/2020 | Population | • African American, American Indian, Asian, Caucasian, Hispanic, Other | 488 (03/08/20) and 459 (09/27/20) individuals, held in facilities outside of NV or in residential confinement, but could not be excluded since based on data provided; NV did not release case data based on: "Under NRS 441A.220, Confidentiality of Information, the release of these 38 offender's [sic] age and race/ethnicity might compromise the medical right to privacy."; DOC reported 0 deaths. |
| NY | 03/01/20201 0/01/2020 | Population | • African American, Asian, Hispanic, Native American, White, Other | Data received 7 months after first filing |
| | 10/01/2020 | Case, death data | • Asian, Black, Hispanic, Native American, White, Other, Unknown | |
| OH | OH DOC consulted a staff attorney in response to our FOIA and did not send updates or data in response to our request. | | | |
| OK | OK DOC indicated they would fulfill our request by 2/12/21 but no data was received | | | |
| OR | 03/01/20201 0/01/2020 | Population | • American Indian, Asian, Black, Hispanic, Pacific Islander, White, Unknown | 0 assumed for "Unknown"; DOC reported they do not track COVID-19 cases by RE |
| | 10/01/2020 | Death data | • Black, Hispanic, White | |
| PA | 03/01/20201 0/01/2020 | Population | • Asian, Black, Hispanic, Indian, White, Other | DOC reported they do not possess records of COVID-19 cases by RE |
| | 10/01/2020 | Death data | • Black, Hispanic, White | 0 deaths assumed for all other RE groups |
| RI | 03/31/2020 09/30/2020 | Population | • American Indian, Asian, Black, Hispanic, White, Other | Cases were not reported by RE because, per DOC, "this is missing for the majority of lab reports."; DOC reported 0 deaths. |
| SC | SC DOC stated they do not track the data we requested. | | | |
| SD | 02/29/2020 09/30/2020 | Population | • Asian, Black, Hispanic or Latino, Native American, Native Hawaiian or Pacific Islander, White, Other | Custody population excludes temporary absences, individuals under SD DOC custody who are housed out of state, and those on extended confinement; DOC reported 0 COVID-19 deaths, RE of positive tests was not recorded |
| TN | FOIA rejected. TN DOC requires a TN resident to file a FOIA. | | | |
| TX | 08/31/2019 | Population | • Black, Hispanic, White, Other and Unknown | Texas Department of Criminal Justice (TDCJ) reported no responsive records and data were confidential; |

| | | | | |
|---|---|---|---|---|
| | | | | Custody population data excluded "Substance Abuse Felony Punishment" facilities |
| | 10/01/2020 | Death data | Black, Hispanic, White, Other | Texas Justice Initiative |
| UT | UT DOC redirected us to their website and did not provide further information. Their website did not contain records responsive to our request. | | | |
| VA | VA DOC requires a VA resident to file a FOIA. | | | |
| VT | 03/01/2020<br>09/30/2020 | Population | • American Indian, Asian/Pacific Islander, Black, Hispanic, Other/Unknown | |
| | 10/15/2020 | Case data | • American Indian/Alaskan Native, Asian/Pacific Islander, Black, White, Other/Unknown | Case data was provided as percentages by RE; 0 deaths from COVID-19 reported |
| WA | 03/31/2020<br>09/30/2020 | Population | • American Indian/Alaska Native, Asian/Pacific Islander, Black, White, Other, Unknown<br>• *Ethnicity categories*: Hispanic Origin | Reported data indicates 1 death (male, race: other), but the DOC website reported 2 deaths (race not reported), our analysis based on data obtained by FOIA |
| | 10/01/2020 | Case data | • Asian/Pacific Islander, Black, American Indian/Alaska Native, White, Other, Unknown | |
| | 10/01/2020 | Death data | • *Race categories:* Other | |
| WI | 02/29/20200<br>9/30/2020 | Population | • American Indian/Alaskan Native, Asian or Pacific Islander, Black, White<br>• *Ethnicity categories*: Hispanic | Custody population by RE was provided as percentages out of a total; Death data not provided due to the following reason: "Our agency does not determine cause or contributing factors of death in any instance. That determination is made by the local medical examiner/coroner. Furthermore, confidentiality laws and privacy protections prevent our agency from sharing any information related to cause of death." |
| | 10/08/2020 | Population | • American Indian/Alaska Native, Asian or Pacific Islander, Black or African American, White, Unknown | |
| WV | 06/2019 | Population | • American Indian/Alaska Native, Asian, Black, Hispanic or Latino, Native Hawaiian or Pacific Islander, White, Multi-Racial or Other | WV Fiscal Year 2019 Annual Report; FOIA required mail submission; Case and death data by RE not reported |
| WY | WY DOC does not have the requested information disaggregated by race/ethnicity. | | | |

All race and ethnicities categories reported below are displayed as reported by their respective state. In our analysis, categories such as "Other," "Multi-Racial," "Two or More" were classified as "Other," while "Unknown" and "Not Specified" were classified as "Unknown."

**Table B. Variation in "Asian American, Native Hawaiian, and Pacific Islander" and "Native American and Alaskan Native" Race Reporting in Custody Population**

| **Race Category Reported** | **States (n=26)** | **No. (%)** |
|---|---|---|
| Asian | CA, CO, CT, ID, KS, MN, NC, NV, NY, PA, RI | 11 (42) |
| Asian American & Pacific Islander | HI, IA, KY, VT, WA, WI | 6 (23) |
| 1) Asian,<br>2) Native Hawaiian/Pacific Islander | MA, MI, SD, WV | 4 (15) |
| 1) Asian<br>2) Pacific Islander | NE, OR | 2 (8) |
| 1) Asian<br>2) Native Hawaiian | ME, ND | 2 (8) |
| None | TX | 1 (4) |
| **Race Category Reported** | **States** | **Freq.** |
| Native American & American Indian | CO, ID, KS, ME, MN, NC, ND, NE, NV, NY, OR, PA, RI, SD, VT | 15 (58) |
| American Indian & Alaskan Native | CT, HI, IA, KY, MA, MI, WA, WI, WV | 9 (35) |
| None | CA, TX | 2 (8) |

### Table C. Race-Disaggregated Crude COVID-19 Case and Death Data by States

| State | Race & Ethnicity | 03.01.20 Population | Cases | Deaths |
|---|---|---|---|---|
| CA | White | 25633 | 3010 (11.7%) | 22 (0.1%) |
| | Black | 35231 | 2707 (7.7%) | 14 (0%) |
| | Hispanic | 55381 | 5478 (9.9%) | 28 (0.1%) |
| | Asian | 1718 | 179 (10.4%) | 4 (0.2%) |
| | Native American | N/A | N/A | 1 |
| | Other | 6396 | 690 (10.8%) | 14 (0.2%) |
| CO | White | 7976 | 415 (5.2%) | 2 (0%) |
| | Black | 3128 | 142 (4.5%) | 0 (0%) |
| | Hispanic | 5588 | 288 (5.2%) | 1 (0%) |
| | Asian | 225 | 10 (4.4%) | 0 (0%) |
| | Native American | 683 | 38 (5.6%) | 0 (0%) |
| | Other | N/A | N/A | N/A |
| CT | White | 3653 | 459 (12.6%) | 1 (0%) |
| | Black | 5349 | 637 (11.9%) | 4 (0.1%) |
| | Hispanic | 3307 | 530 (16%) | 2 (0.1%) |
| | Asian | 61 | 4 (6.6%) | 0 (0%) |
| | Native American | 41 | 4 (9.8%) | 0 (0%) |
| | Other | N/A | N/A | N/A |
| HI | White | 1266 | N/A | 0 (0%) |
| | Black | 230 | N/A | 0 (0%) |
| | Hispanic | N/A | N/A | N/A |
| | Asian | 3397 | N/A | 0 (0%) |
| | Native American | 53 | N/A | 0 (0%) |
| | Other | 326 | N/A | 0 (0%) |
| IA | White | 5561 | 798 (14.3%) | 3 (0.1%) |
| | Black | 2128 | 269 (12.6%) | 1 (0%) |
| | Hispanic | 580 | 60 (10.3%) | 0 (0%) |
| | Asian | 68 | 13 (19.1%) | 0 (0%) |
| | Native American | 167 | 22 (13.2%) | 0 (0%) |
| | Other | N/A | N/A | N/A |
| ID | White | 5854 | 1216 (20.8%) | 2 (0%) |
| | Black | 260 | 57 (21.9%) | 0 (0%) |
| | Hispanic | 1154 | 266 (23.1%) | 0 (0%) |
| | Asian | 34 | 6 (17.6%) | 0 (0%) |

| State | Race & Ethnicity | 03.01.20 Population | Cases | Deaths |
|---|---|---|---|---|
| | Native American | 304 | 80 (26.3%) | 0 (0%) |
| | Other | 213 | 27 (22.3%) | 0 (0%) |
| KS | White | 6852 | N/A | 2 (0%) |
| | Black | 2828 | N/A | 5 (0.2%) |
| | Hispanic | 1235 | N/A | 0 (0%) |
| | Asian | 94 | N/A | 0 (0%) |
| | Native American | 270 | N/A | 0 (0%) |
| | Other | N/A | N/A | N/A |
| KY | White | 8639 | N/A | 6 (0.1%) |
| | Black | 3125 | N/A | 5 (0.2%) |
| | Hispanic | 232 | N/A | 0 (0%) |
| | Asian | 26 | N/A | 0 (0%) |
| | Native American | 10 | N/A | 0 (0%) |
| | Other | 170 | N/A | N/A |
| MA | White | 3334 | N/A | 4 (0.1%) |
| | Black | 2259 | N/A | 2 (0.1%) |
| | Hispanic | 2064 | N/A | 0 (0%) |
| | Asian | 119 | N/A | 0 (0%) |
| | Native American | 44 | N/A | 1 (2.3%) |
| | Other | 277 | N/A | 1 (0.9%) |
| ME | White | 1732 | N/A | 0 (0%) |
| | Black | 225.6 | N/A | 0 (0%) |
| | Hispanic | N/A | N/A | N/A |
| | Asian | 20.38 | N/A | 0 (0%) |
| | Native American | 56.8 | N/A | 0 (0%) |
| | Other | N/A | N/A | N/A |
| MI | White | 16545 | 3239 (19.6%) | 36 (0.2%) |
| | Black | 19971 | 3682 (18.4%) | 33 (0.2%) |
| | Hispanic | 998 | 192 (19.2%) | 1 (0.1%) |
| | Asian | 133 | 20 (15%) | 0 (0%) |
| | Native American | 422 | 88 (20.9%) | 0 (0%) |
| | Other | N/A | N/A | N/A |
| MN | White | 4101 | 227 (5.5%) | 1 (0%) |
| | Black | 3317 | 154 (4.6%) | 1 (0%) |
| | Hispanic | 489 | 30 (6.1%) | 0 (0%) |
| | Asian | 235 | 10 (4.3%) | 0 (0%) |
| | Native American | 778 | 39 (5%) | 0 (0%) |

| State | Race & Ethnicity | 03.01.20 Population | Cases | Deaths |
|---|---|---|---|---|
| | Other | 9 | N/A | N/A |
| NC | White | 13904 | 1266 (9.1%) | 6 (0%) |
| | Black | 17805 | 1471 (8.3%) | 9 (0.1%) |
| | Hispanic | 1884 | 259 (13.7%) | 0 (0%) |
| | Asian | 102 | 11 (10.8%) | 0 (0%) |
| | Native American | 681 | 66 (9.7%) | 0 (0%) |
| | Other | 207 | 24 (11.6%) | 1 (0.1%) |
| ND | White | 1092 | 17 (1.6%) | 0 (0%) |
| | Black | 190 | 2 (1.1%) | 0 (0%) |
| | Hispanic | 112 | 0 (0%) | 0 (0%) |
| | Asian | 10 | 0 (0%) | 0 (0%) |
| | Native American | 391 | 7 (1.8%) | 0 (0%) |
| | Other | 11 | 0 (0%) | 0 (0%) |
| NE | White | 2949 | 132 (4.5%) | 0 (0%) |
| | Black | 1555 | 64 (4.1%) | 0 (0%) |
| | Hispanic | 822 | 37 (4.5%) | 0 (0%) |
| | Asian | 52 | 1 (1.9%) | 0 (0%) |
| | Native American | 276 | 5 (1.8%) | 0 (0%) |
| | Other | 38 | 2 (5.3%) | 0 (0%) |
| NV | White | 5545 | N/A | 0 (0%) |
| | Black | 3954 | N/A | 0 (0%) |
| | Hispanic | 2718 | N/A | 0 (0%) |
| | Asian | 370 | N/A | 0 (0%) |
| | Native American | 227 | N/A | 0 (0%) |
| | Other | 58 | N/A | 0 (0%) |
| NY | White | 10486 | 127 (1.2%) | 1 (0%) |
| | Black | 21251 | 391 (1.8%) | 10 (0%) |
| | Hispanic | 10624 | 236 (2.2%) | 4 (0%) |
| | Asian | 256 | 6 (2.3%) | 2 (0.8%) |
| | Native American | 408 | 9 (2.2%) | 0 (0%) |
| | Other | 761 | 10 (1.3%) | 0 (0%) |
| OR | White | 10370 | N/A | 6 (0.1%) |
| | Black | 1382 | N/A | 1 (0.1%) |
| | Hispanic | 1948 | N/A | 2 (0.1%) |
| | Asian | 263 | N/A | 0 (0%) |
| | Native American | 469 | N/A | 0 (0%) |
| | Other | 3 | N/A | N/A |

| State | Race & Ethnicity | 03.01.20 Population | Cases | Deaths |
|---|---|---|---|---|
| PA | White | 19431 | N/A | 3 (0%) |
| | Black | 20631 | N/A | 7 (0%) |
| | Hispanic | 4313 | N/A | 1 (0%) |
| | Asian | 118 | N/A | 0 (0%) |
| | Native American | 40 | N/A | 0 (0%) |
| | Other | 210 | N/A | 0 (0%) |
| RI | White | 961 | N/A | 0 (0%) |
| | Black | 707 | N/A | 0 (0%) |
| | Hispanic | 646 | N/A | 0 (0%) |
| | Asian | 38 | N/A | 0 (0%) |
| | Native American | 24 | N/A | 0 (0%) |
| | Other | 46 | N/A | 0 (0%) |
| SD | White | 1980 | N/A | 0 (0%) |
| | Black | 309 | N/A | 0 (0%) |
| | Hispanic | 149 | N/A | 0 (0%) |
| | Asian | 27 | N/A | 0 (0%) |
| | Native American | 1305 | N/A | 0 (0%) |
| | Other | 11 | N/A | 0 (0%) |
| TX | White | 47191 | N/A | 60 (0.1%) |
| | Black | 45761 | N/A | 64 (0.1%) |
| | Hispanic | 46375 | N/A | 73 (0.2%) |
| | Asian | N/A | N/A | N/A |
| | Native American | N/A | N/A | N/A |
| | Other | 797 | N/A | 2 (0.3%) |
| VT | White | 1402 | 44 (3.1%) | 0 (0%) |
| | Black | 153 | 9 (5.9%) | 0 (0%) |
| | Hispanic | 4 | 0 (0%) | 0 (0%) |
| | Asian | 6 | 0 (0%) | 0 (0%) |
| | Native American | 18 | 0 (0%) | 0 (0%) |
| | Other | 56 | 2 (3.6%) | 0 (0%) |
| WA | White | 13083 | 325 (2.5%) | 0 (0%) |
| | Black | 3308 | 68 (2.1%) | 0 (0%) |
| | Hispanic | 2726 | 71 (2.6%) | N/A |
| | Asian | 808 | 16 (2%) | 0 (0%) |
| | Native American | 1109 | 34 (3.1%) | 0 (0%) |
| | Other | 300 | 8 (2.7%) | 1 (0.3%) |
| WI | White | 12142 | 662 (5.5%) | N/A |

| State | Race & Ethnicity | 03.01.20 Population | Cases | Deaths |
|---|---|---|---|---|
| | Black | 9902 | 763 (7.7%) | N/A |
| | Hispanic | 1785 | N/A | N/A |
| | Asian | 232 | 20 (8.6%) | N/A |
| | Native American | 989 | 49 (5%) | N/A |
| | Other | N/A | N/A | N/A |
| WV | White | 5012 | 0 (0%) | 0 (0%) |
| | Black | 727 | 0 (0%) | 0 (0%) |
| | Hispanic | 25 | 0 (0%) | 0 (0%) |
| | Asian | 5 | 0 (0%) | 0 (0%) |
| | Native American | 4 | 0 (0%) | 0 (0%) |
| | Other | 48 | N/A | 3 (6.2%) |
